# Opposing and antagonizing effects of SARS-CoV-2/COVID-19 infection and recombinant zoster vaccination on the risk of late-onset Alzheimer disease

**DOI:** 10.64898/2026.01.21.26344555

**Authors:** Carly M. Rose, Shiying Liu, William S. Bush, Jonathan L. Haines, Scott M. Williams, Dana C. Crawford, N3C Consortium

## Abstract

Initial data indicate SARS-CoV-2 and herpes zoster negatively impact cognition, but studies examining their independent or synergistic impact specifically for late-onset Alzheimer disease (LOAD) risk are lacking. Retrospective cohort analysis of older individuals (age ≥65 between January 20, 2020 and May 16, 2025; n∼1.5 million) from the U.S. National Clinical Cohort Collaborative (N3C) COVID-19 Enclave indicates that, in adjusted Cox proportional hazard models, those with ≥1 reported COVID-19 infection were at increased LOAD risk compared with those with no reported infection (HR=1.13, 95% CI: 1.10-1.16). Recombinant zoster vaccinated patients had decreased LOAD risk regardless of sex, race, age at baseline and number of clinic visits (HR=0.71, 95% CI: 0.69-0.74); however, COVID-19 infection mitigated the protective effect of the vaccination. Recombinant zoster vaccination and reduced exposure to COVID-19 infection in the later decades of life may reduce the risk of developing Alzheimer disease over at least a five-year period.

---

Alzheimer disease (AD) is a progressive neurodegenerative disorder characterized by memory loss, cognitive decline, and behavioral changes that impact daily functioning^1^. Late-onset Alzheimer disease (LOAD; onset age ≥65)^2^, currently affects an estimated 7.2 million Americans and is expected to increase to 13.8 million by 2060^3^. Known risk factors include non-modifiable factors such as advanced age, female sex, and genetics (*APOE*)^4,5^. Modifiable risk factors include education^6^ and physical activity^7^. Recent evidence also indicates that both the live-attenuated (Zostavax) and the newer recombinant zoster (Shingrix) vaccines reduce dementia risk^8–10^ while herpes zoster infection (also known as shingles) caused by varicella-zoster virus reemergence increases dementia risk^11^.

Several studies have highlighted associations between viral infections and increased neurodegeneration risk^12^, indicating infectious diseases as potential modifiable risk factors that evoke the adaptive immune system, resulting in possibly damaging neuroinflammation. The Coronavirus disease 2019 (COVID-19) results from a highly infectious coronavirus that quickly developed into a global pandemic resulting in substantial morbidity (e.g., long COVID^13^) and mortality^14^ in the United States. Early in the pandemic, COVID-19’s triggering of temporary anosmia and reports of “brain fog” indicated its infection has a neurological impact^15^. Pre-COVID and post-COVID infection magnetic resonance imaging and cognitive studies in the UK Biobank demonstrated adverse brain changes and greater average cognitive decline between the time-points^16^. The Atherosclerosis Risk in Communities (ARIC) study also demonstrated a decrease in cognitive function among those hospitalized for COVID-19 compared with those not infected with SARS-CoV-2^17^, and at least one small prospective cohort recently demonstrated an increase in incident AD-type mild cognitive impairment among those with long COVID compared with recovered COVID or COVID-negativity^18^.

The National Clinical Cohort Collaborative (N3C) COVID-19 Enclave, a U.S. resource of harmonized patient-level data aggregated in response to the pandemic^19^, presents a unique resource to directly investigate possible associations between COVID-19 infection and LOAD alone or in combination with Shingrix vaccination status using large-scale, multi-institutional electronic health record data.

## Online Methods

The N3C data transfer from healthcare Institutions to National Center for Advancing Translational Sciences (NCATS) was performed under a Johns Hopkins University Reliance Protocol # IRB00249128 or individual site agreements with NIH. The N3C Data Enclave is managed under the authority of the NIH; information can be found at https://ncats.nih.gov/n3c/resources.

### Study Population

We accessed U.S. multi-institutional healthcare data from the N3C COVID-19 Enclave comprising electronic health records (EHRs) using the Limited Data Set (Level 2 access) for over 24 million patients with over 36 billion medical records (v192). Using these data, we conducted a retrospective cohort study anchored to January 20, 2020, when the CDC reported the first laboratory-confirmed case of the 2019 novel coronavirus in the US^20^. The data transfer agreements (DTAs) executed between institutions and NCATS for contributions to the N3C COVID Enclave were initially limited to a five-year term, which began to expire in May 2025^21^. Our study period was January 20, 2020, to May 16, 2025, concordant with the final data release, creating at maximum an ∼5-year follow-up window. Patients were excluded if they had a zero-visit history in the defined baseline period (January 20, 2019, and January 20, 2020), had a LOAD diagnosis before the study initiation date (January 20, 2020), and/or were under the age of 65 (**Supplementary Figure 1**).

### Phenotyping

The N3C defines a COVID-19 confirmed patient as meeting one of the following; 1) having one or more COVID-19 positive laboratory result(s), 2) having one or more strong positive diagnosis code, or 3) having two or more “weak positive” diagnosis codes during the same encounter or on the same date on or prior to May 1, 2020^22,23^. Each COVID-19-positive patient was matched to approximately two control patients, which the N3C defines as a patient with one or more of the lab tests in the labs table with a non-positive result^22^. The number of COVID-19 diagnoses was defined according to the N3C COVID-19 confirmed patient phenotyping guidelines and the CDC-defined guidelines with reinfection being at least 90 days between infections^24,25^. Patients were categorized into none, one, and two or more infections. We defined LOAD using the Dementia Tenant Alzheimer’s concept set, a curated list of 40 concept IDs developed by N3C Logic Liaison teams and clinical experts^26,27^. Given our interest in LOAD specifically, this concept set was further refined to exclude the concept condition names that included a mention of “early onset”. Shingrix vaccination status was defined using the drug exposure Observational Medical Outcomes Partnership (OMOP) table to include patients with a drug concept name “Shingrix” or a combination of “zoster” and “recombinant” in that field. Clinic visits are defined by aggregating individual visit records to the patient level and counting the number of unique visit dates per patient. Demographic variables including age, sex, and race were defined from the “person” OMOP table, while age was further calculated to be the difference between birth dates and baseline date (1/20/2020) to produce “age at baseline”. The race variable included “White”, “Black or African American”, “Asian”, “American Indian or Alaska Native”, “Native Hawaiian or Other Pacific Islander”, and “Other/Unknown”. Individuals with “Hispanic” listed as race were excluded due to entry errors in the race category. To understand general health behaviors, we captured information on history of mammography, colonoscopy, and prostate screenings. Mammography and colonoscopy information was captured in the procedure table to include patients who have “screening mammography” and “screening colonoscopy” concept names, respectively, while prostate-specific antigen was found in the measurement table using “prostate specific antigen”, or “PSA” concept names. We also defined a positive pneumococcal vaccine status as those who have a drug concept name “pneumococcal” using the drug exposure OMOP table.

### Statistical Methods

Propensity score matching was performed using the *matchit* R package and nearest neighbor methods to ensure COVID-19-confirmed cases and non-cases have similar distributions based on age, sex, and race. The effectiveness of propensity score matching was assessed using a Love plot (**Supplementary Figure 2**). To estimate associations between LOAD (outcome), COVID-19 (exposure), and Shingrix (exposure), both independently and in combination, we conducted univariate and multivariate logistic regressions and survival analyses. The univariate models included associating COVID-19 and LOAD (outcome), as well as Shingrix and LOAD (outcome), with both models adjusted for age at baseline, sex, race, and number of clinic visits. Time-fixed and time-varying Cox proportional hazards models were used for survival analysis models with the primary outcome of LOAD and time-to-event definitions, including time to LOAD diagnosis for LOAD patients and time to last visit date for non-LOAD patients. Time-varying models also defined exposures of interest (reported COVID-19 infection and Shingrix vaccination, respectively) based on individual patient dates of reported infection or vaccination, and were created using the *tmerge* function from the R *survival* package. Each patient’s observation period was split into sequential intervals at the dates of COVID-19 infection and/or Shingrix vaccination, such that the covariate status of each exposure reflects the patient’s true exposure state within each interval. Cox proportional hazards models were then fit using the counting process formulation *Surv(tstart, tstop, event),* which handles the interval structure produced by *tmerge* and allows both exposures to serve simultaneously as time-varying covariates within the same model.

To ensure analyses focused on incident LOAD cases following COVID-19 and Shingrix exposure respectively, we excluded patients with LOAD diagnosis prior to their first confirmed COVID-19 infection or Shingrix vaccination, respectively. This allowed us to assess potential independent temporal relationships between COVID-19 and Shingrix exposures with incident LOAD diagnosis. To capture uncertainty of COVID-19 case status and timing of infection as well as timing of Shingrix vaccination, sensitivity analyses included a combined model of COVID-19 and Shingrix any time during the follow up period with LOAD (outcome), unadjusted and adjusted for age at baseline, sex, race, and number of clinic visits as well as a full model that included an interaction term between COVID-19 and Shingrix. All analyses were done within the N3C cloud environment using SparkR (version 4.3.3) and PySpark (version 3.9.23).

## Results

From a final cohort of approximately 1.5 million patients (**Supplementary Figure 1**), 40,048 were diagnosed with incident LOAD. Compared with non-LOAD patients (n=1,439,532), LOAD patients were on average older, and more likely to be female and Black (p<0.001; **Table 1**). The proportion of Shingrix vaccination was lower among incident LOAD patients compared to non-LOAD patients (19.4% vs. 15.2%), and among those vaccinated, the average age at vaccination was higher among LOAD patients (75.6 years) compared to non-LOAD patients (71.9 years; p<0.001; **Table 1**).

**Table 1.** N3C patient distribution ≥65 years, stratified by LOAD status, propensity score matched on confirmed COVID-19 infection.

|  | **Non-LOAD**<br>**(n=1,479,500)** | **LOAD**<br>**(n=40,048)** | **P-value** |
| --- | --- | --- | --- |
| **Age at Baseline (Jan 20, 2020), mean (SD)** | 72.1 (5.1) | 75.7 (4.9) | <0.001 |
| **Age at First Shingrix Vaccination, mean (SD)** | 71.9 (5.0) | 75.6 (5.1) | <0.001 |
| **Female Sex, n (%)** | 796,664 (55.3%) | 23,684 (59.1%) | <0.001 |
| **Race, n (%)** |  |  |  |
| *White* | 1,139,099 (79.1%) | 30,078 (75.1%) | <0.001 |
| *Black* | 139,122 (9.7%) | 5,079 (12.7%) |  |
| *Asian* | 38,741 (2.7%) | 1,241 (3.1%) |  |
| *Native Hawaiian or Other Pacific Islander* | 3,476 (0.2%) | 150 (0.4%) |  |
| *American Indian or Alaska Native* | 5,417 (0.4%) | 185 (0.5%) |  |
| *Other/Unknown* | 113,677 (7.9%) | 3,315 (8.3%) |  |
| **Had COVID-19, n (%)** | 718,501 (49.9%) | 21,248 (53.1%) | <0.001 |
| **Total Clinic Visits, mean (SD)** | 114.3 (110.6) | 119.2 (118.0) | <0.001 |
| **Had Shingrix Vaccine, n (%)** | 279,452 (19.4%) | 6,094 (15.2%) | <0.001 |
| **Had Pneumococcal Vaccine, n (%)** | 410,341 (28.5%) | 10,067 (25.1%) | <0.001 |

N3C = National Clinical Cohort Collaborative; LOAD = Late-onset Alzheimer's Disease; PSM = Propensity Score Matching; SD = standard deviation. Statistical comparisons between LOAD and non-LOAD groups were performed using chi-square tests for categorical variables and t-tests for continuous variables. P-values <0.05 were considered statistically significant.

We first directly compared LOAD risk between COVID-19 positive and negative individuals and whether these associations were modified by reported Shingrix vaccination status. In time-fixed Cox proportional hazards model, where first COVID-19 infection date is the baseline for COVID-19-positive patients and COVID-19-absent patients used the baseline date January 20, 2020 (**Methods**), COVID-19 infection was associated with a 13% increased hazard of incident LOAD (HR=1.13, 95% CI: 1.10-1.16, p<2.0x10^-16^) adjusting for age at baseline, sex, race, and number of clinic visits. The addition of Shingrix vaccination status to the model yielded a very similar COVID-19 infection hazard estimate of LOAD risk (HR=1.14, 95% CI: 1.11-1.16, p<2.0x10^-16^), suggesting association between COVID-19 infection and LOAD risk is independent of Shingrix vaccination status. COVID-19 time-varying Cox proportional hazards models yielded similar results: COVID-19 infection was associated with a 15% increased hazard of incident LOAD after adjustment for age, sex, race, and number of clinic visits (HR=1.15, 95% CI: 1.12-1.18, p<2.0x10^-16^). The addition of Shingrix vaccination to the COVID-19 time-varying model did not change the risk of incident LOAD with COVID-19 infection (HR=1.15, 95% CI: 1.12-1.18, p<2.0x10^-16^). Lastly, treating both COVID-19 infection and Shingrix vaccination as time-varying exposures in the fully adjusted model produced identical results as just the COVID-19 time-varying model.

We then made similar comparisons between risk of incident LOAD and reported Shingrix vaccination status. In a time-fixed Cox proportional hazards model adjusted for age at baseline, sex, race, and number of clinic visits, Shingrix vaccination was associated with a 29% reduced hazard of LOAD (HR=0.71, 95% CI: 0.69-0.74, p<2.0x10^-16^), and the addition of reported COVID-19 infection to the model did not change the reduced hazard of incident LOAD associated with Shingrix vaccination (HR=0.71, 95% CI: 0.69-0.74, p<2.0x10^-16^). The Shingrix time-varying model with the same adjustments reported nearly identical results (HR=0.72, 95% CI: 0.70-0.74, p<2.0x10^-16^), with the addition of COVID-19 infection not changing the effect. The model with both Shingrix and COVID-19 treated as time-varying exposures also produced similar results, with a 30% reduced hazard of incident LOAD (HR=0.70, 95% CI: 0.67-0.72, p<2.0x10^-16^), a result that remained unchanged with the addition of COVID-19 as an exposure.

We then investigated whether there was statistical evidence that COVID-19 infection and Shingrix vaccination, both independent associations in the opposite direction, might exert an antagonistic joint effect on incident LOAD risk. To do this, we first assessed a time-fixed Cox proportional hazards model with incident LOAD (outcome), COVID-19 infection (exposure), and Shingrix vaccination (exposure), adjusting for age at baseline, sex, race, and number of clinic visits. With the addition of an interaction term between COVID-19 infection and Shingrix vaccination to the fully adjusted model, the effects of each were similar to the individual models (COVID-19 HR: 1.11, 95% CI: 1.08-1.14, p<2.0x10^-16^; Shingrix HR: 0.67, 95% CI: 0.64-0.70, p<8.0x10^-13^), with the interaction term HR of 1.18 (95% CI: 1.10-1.26, p<1.04x10^-6^). Results from COVID-19 time-varying Cox proportional hazard models for the fully adjusted model were similar: COVID-19 HR=1.14, 95% CI: 1.11-1.17, p<2.0x10^-16^; Shingrix HR=0.65, 95% CI: 0.63-0.67, p<2.0x10^-16^, and interaction term HR=1.29, 95% CI: 1.21-1.38, p<2.3x10^-14^.

To examine the possible antagonism between Shingrix vaccination status with respect to incident LOAD risk further, we stratified by Shingrix vaccination status and used the same baseline dates as above for time-fixed Cox proportional hazards modeling with COVID-19 infection as the exposure. Among Shingrix vaccinated patients, adjusted for age at baseline, sex, and number of clinic visits, COVID-19 infection had a 31% increased hazard of LOAD (HR=1.31, 95% CI: 1.22-1.39, p<1.4x10^-15^), indicating COVID-19 infection negates the protective effect on incident LOAD. Among patients without evidence of Shingrix vaccination, COVID-19 was associated with an 11% hazard of LOAD (HR=1.11, 95% CI: 1.08-1.14, p<9.5x10^-14^). COVID-19 time-varying Cox proportional hazards models yielded similar results, showing COVID-19 had a 25% increased hazard of incident LOAD among Shingrix vaccinated patients (HR=1.25, 95% CI: 1.17-1.34, p<1.1x10^-11^), and a 12% increased hazard among patients without evidence of Shingrix vaccination (HR=1.12, 95% CI: 1.09-1.16, p<3.5x10^-16^).

We also stratified by COVID-19 infection status and used the same baseline dates as above for time-fixed Cox proportional hazards modeling with Shingrix vaccination as the exposure (**Figure 1 and Table 2**). Among COVID-19 patients, adjusted for age at baseline, sex, race, and number of clinic visits, Shingrix vaccination had a 26% reduced hazard of incident LOAD (HR=0.74, 95% CI: 0.70-0.78, p<5.0x10^-30^). Among COVID-19-absent individuals, Shingrix vaccination was associated with a 30% reduced hazard of incident LOAD (HR=0.70, 95% CI: 0.67-0.73, p<2.0x10^-62^), indicating Shingrix vaccination reduces LOAD risk regardless of COVID-19 infection status. Shingrix vaccination time-varying Cox proportional hazards models yielded similar results: among COVID-19 patients, Shingrix vaccination was associated with a 31% reduced hazard of incident LOAD after adjustment for age, sex, race, and number of clinic visits (HR=0.69, 95% CI: 0.65-0.73, p<2.0x10^-16^), while among COVID-19-absent patients, Shingrix vaccination had a 28% reduced hazard of incident LOAD (HR=0.72, 95% CI: 0.68-0.75, p<2.0x10^-16^).

**Figure 1.**
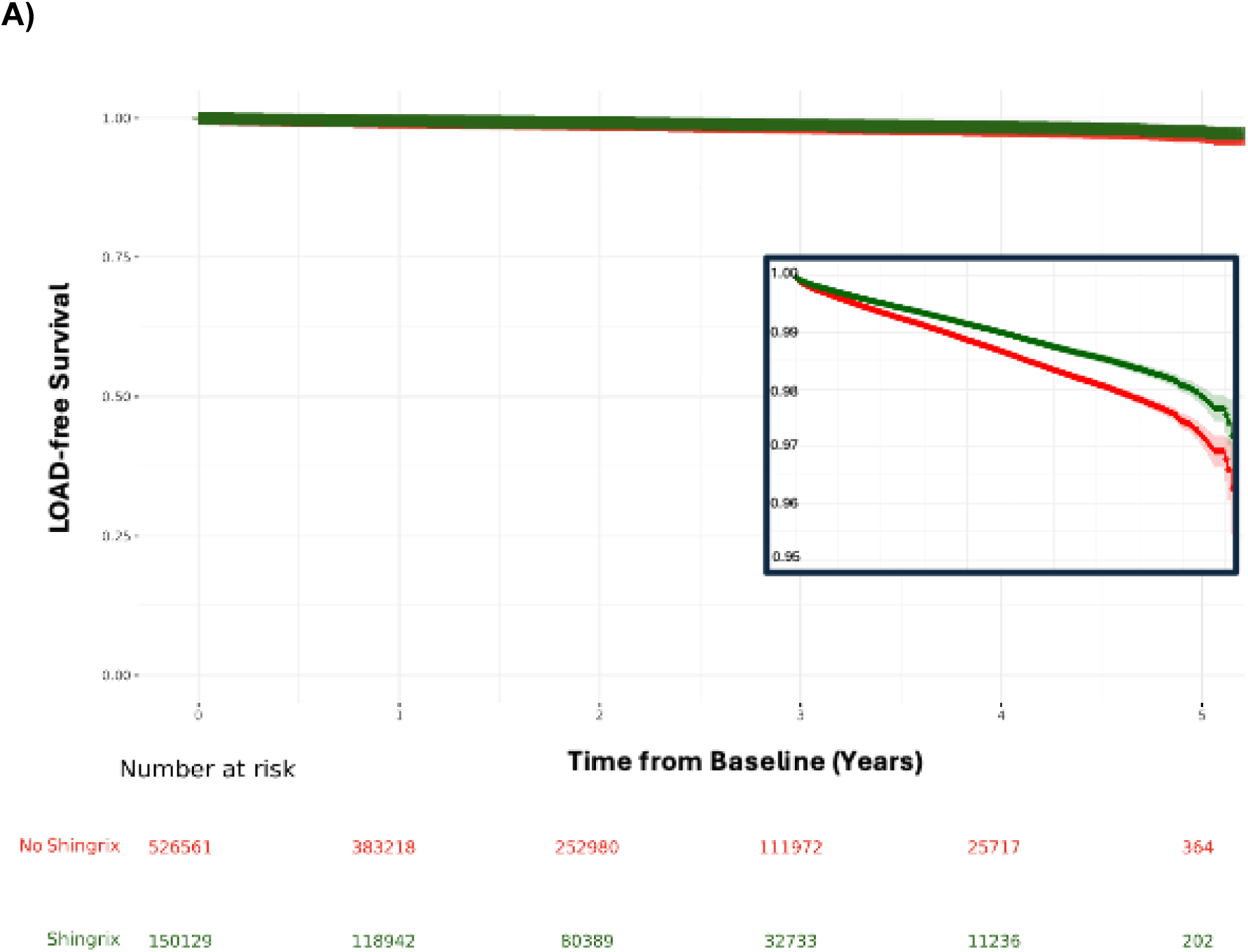
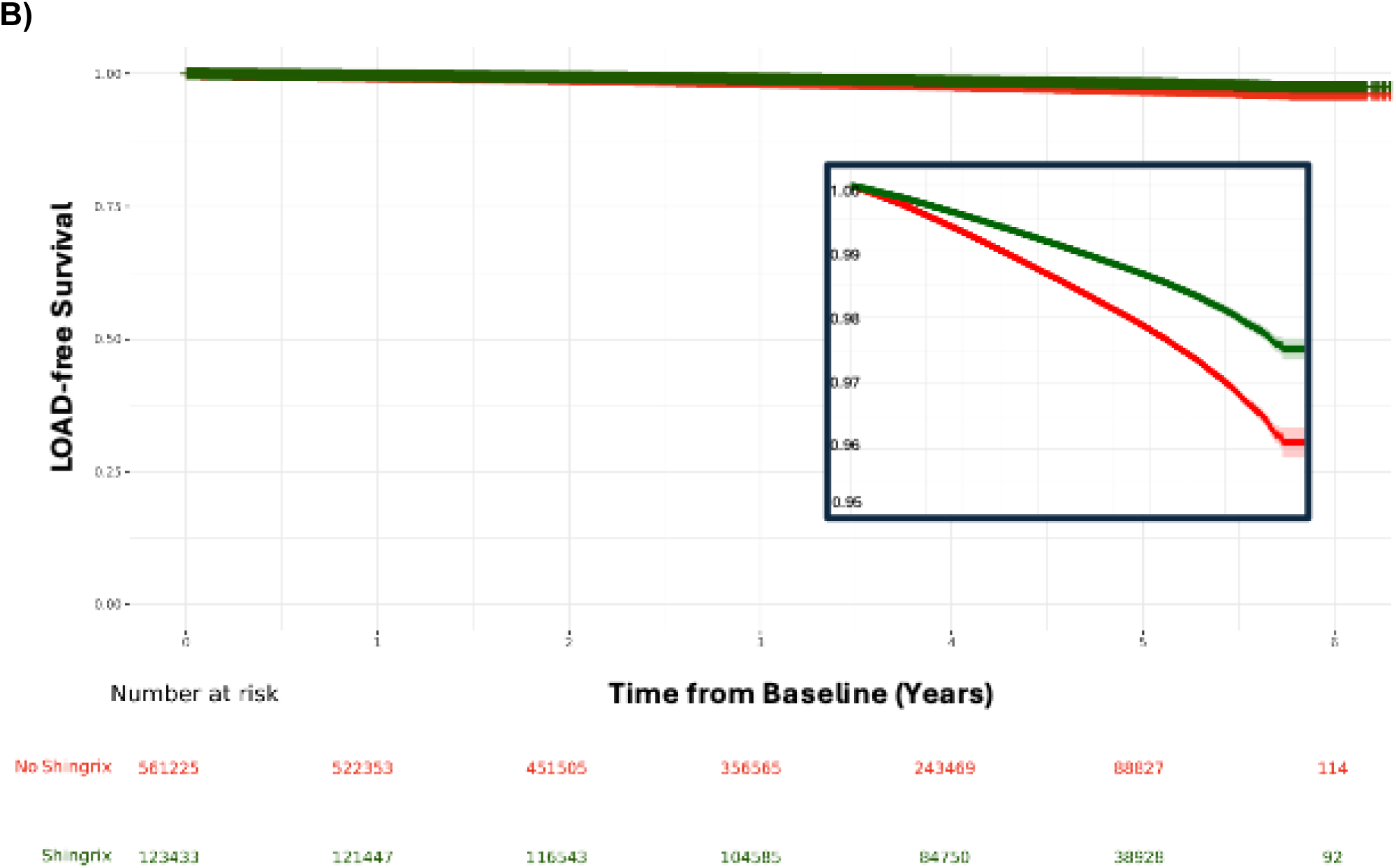
Cox-proportional hazard model for late-onset Alzheimer disease-free survival among a) COVID-19 patients and b) COVID-19-free patients ≥65 years of age by Shingrix vaccination status. Individual first infection date for COVID-19 patients (n=676,690) was the baseline date whereas January 20,2020 was the baseline date for COVID-19-free patients (n=684,658). The censor date is incident late-onset Alzheimer disease (LOAD; y-axis) or last clinic visit date in follow-up period among COVID-19 patients (panel A; n=10,102) and COVID-19-free patients (panel B; n=18,792) during the follow-up period (x-axis) by Shingrix vaccination status (red=yes and green=no). Models were adjusted for age, sex, race, and number of clinic visits. LOAD-free survival curves are shown with 95% confidence intervals (shaded regions). Numbers at risk estimated using hazard ratios from Cox proportional hazards model, with baseline sample sizes, event rates, and median follow-up from observed data.

**Table 2.** Cox-proportional hazard ratios for late-onset Alzheimer disease-free survival among COVID-19 patients and COVID-19-free patients ≥65 years of age by Shingrix vaccination status. Unadjusted and adjusted for age, sex, race, and number of clinic visits.

|  | **Shingrix**<br>**vaccination**<br>**status (incident**<br>**LOAD events)** | **Unadjusted HR**<br>**(95% CI)** | **P-value** | **Adjusted HR**<br>**(95% CI)** | **P-value** |
| --- | --- | --- | --- | --- | --- |
| **COVID-19**<br>**patients**<br>**(n=676,690)** | n<sub>yes</sub> = 150,129<br>(n= 1,782)<br>n<sub>no</sub> = 526,561<br>(n= 8,320) | 0.697<br>(0.662-0.734) | 2.24x10<sup>-43</sup> | 0.738<br>(0.700-0.777) | 4.46x10<sup>-30</sup> |
| **COVID-19-free**<br>**patients**<br>**(n=684,658)** | n<sub>yes</sub> = 123,433<br>(n= 2,651)<br>n<sub>no</sub> = 561,225<br>(n= 16,141) | 0.584<br>(0.561-0.609) | 2.79x10<sup>-144</sup> | 0.698<br>(0.670-0.728) | 1.44x10<sup>-62</sup> |

LOAD = Late-onset Alzheimer's Disease; HR = Hazard ratio; CI = Confidence interval. Statistical comparisons between LOAD and non-LOAD groups were performed using chi-square tests for categorical variables and t-tests for continuous variable.

We conducted several sensitivity analyses to test the robustness of our findings, including logistic regression which treats LOAD as a binary outcome regardless of time to diagnosis. Results of these analyses were consistent with our primary findings and showed that having at least one reported COVID-19 infection compared to those without infection associated with higher odds of LOAD adjusted for age at baseline, sex, race, and number of clinic visits (OR=1.13, 95% CI: 1.10-1.15, p<3.0x10^-30^) (**Figure 2a**). Conversely, patients with Shingrix vaccination had lower odds of LOAD compared with those without evidence of the vaccination (OR=0.78, 95% CI: 0.75-0.80, p<3.0x10^-68^), also adjusted for age at baseline, sex, race, and number of clinic visits (**Figure 2b**). Compared to the univariate Shingrix result (OR=0.75, 95% CI: 0.72-0.77, p<2x10^-16^), modeling one covariate at a time showed age at baseline had the greatest influence on the impact of Shingrix vaccination on LOAD (OR=0.81, 95% CI: 0.79-0.83, p<2x10^-16^), while sex (OR=0.74, 95% CI: 0.72-0.76, p<2x10^-16^) and race (OR=0.75, 95% CI: 0.73-0.77, p<2x10^16^) did not influence the effect, and the number of clinic visits marginally increased the effect (OR=0.71, 95% CI: 0.69-0.73, p<2x10^-16^. (**Figure 3**).

**Figure 2.**
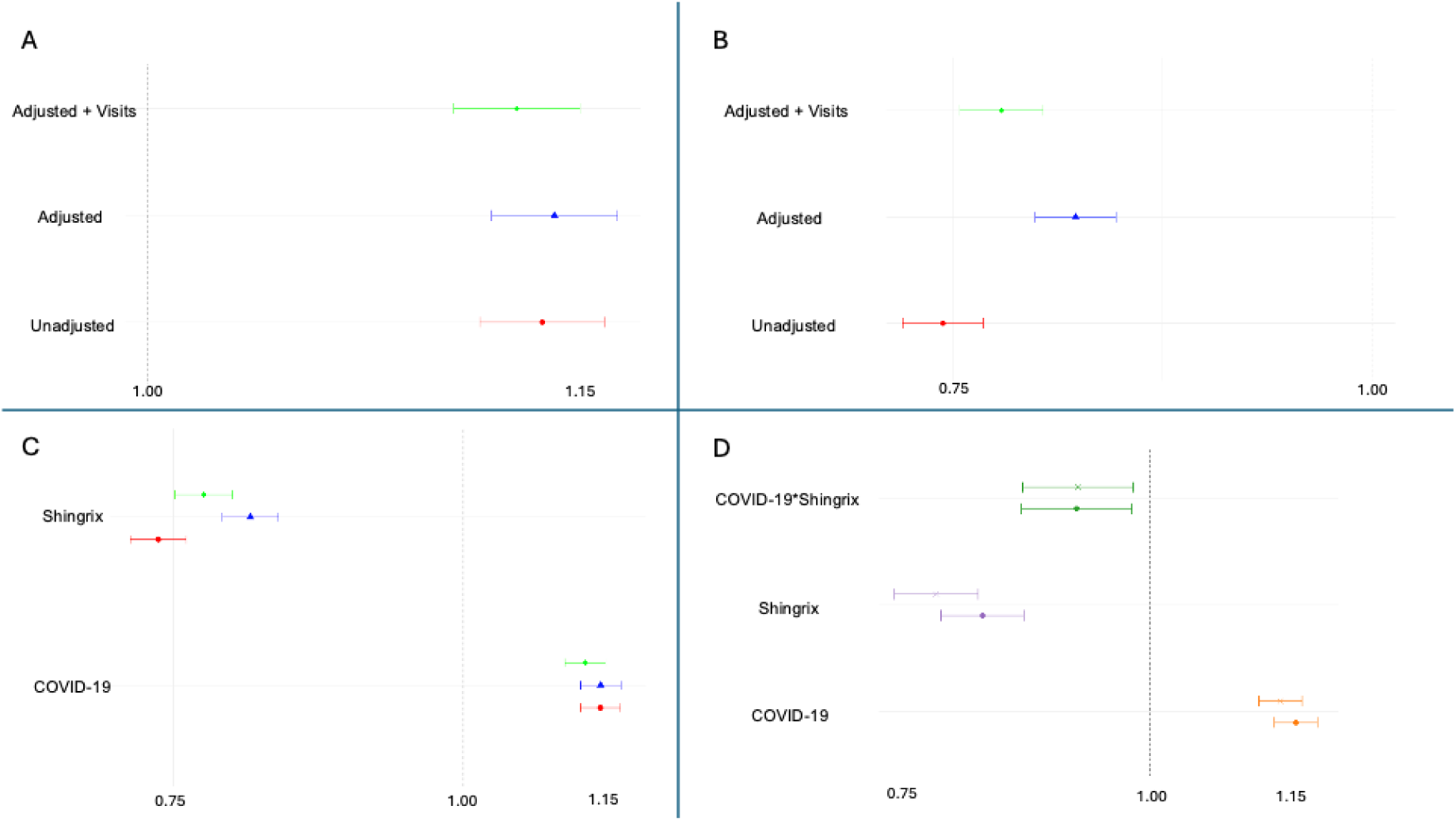
Association between COVID-19 infection and Shingrix vaccination status and late-onset Alzheimer disease among patients ≥65 years of age. Patients were propensity score matched on confirmed COVID-19 infection as described in Methods. A series of logistic regression models focused on late-onset Alzheimer disease (LOAD) anytime during the follow-up period (outcome) and (a) COVID-19 infection only, (b) Shingrix vaccination status only, (c) COVID-19 infection and Shingrix vaccination status, and (d) COVID-19 infection, Shingrix vaccination status, and interaction term. Unadjusted (red) and adjusted for age at baseline, sex, race (blue), and adjusted with the addition of the number of clinic visits (green) odds ratios and 95% confidence intervals are plotted on the x-axis for panels A-C. For panel D, the adjusted odds ratios and 95% confidence intervals from the full logistic regression model are shown for COVID-19 infection (orange), Shingrix vaccination (purple), and the interaction term for COVID-19 infection and Shingrix vaccination status. The circles for each term represent the fully adjusted model with COVID-19 infection, Shingrix vaccination, age, sex, race, and the interaction term, where the stars represent the addition of the number of clinic visits in the model.

**Figure 3.**
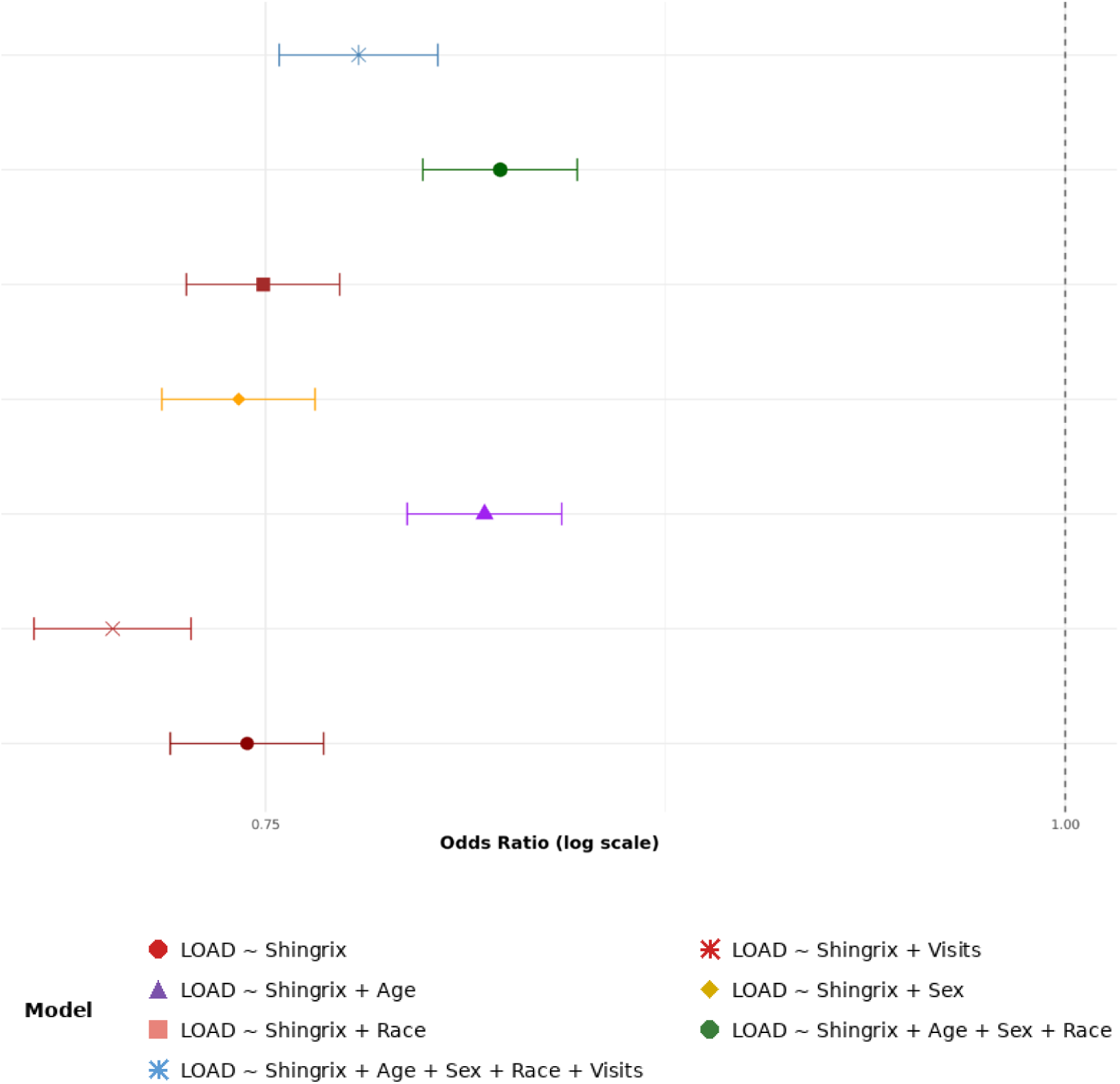
Covariate progressive addition models for association between Shingrix vaccination status and late-onset Alzheimer disease among patients ≥65 years of age. Patients were propensity score-matched by COVID-19 infection as detailed in Methods. Plotted are odds ratios and 95% confidence intervals for various logistic regression models for late-onset Alzheimer disease (LOAD) and Shingrix vaccination status. Starting at the bottom, the basic model of exposure (Shingrix vaccination status) and LOAD (outcome) is shown followed by the model with Shingrix vaccination status and age at baseline (purple); Shingrix vaccination status, age at baseline, sex, and race (green); Shingrix vaccination status and race (red); and Shingrix vaccination status and sex (yellow).

Sensitivity analyses using logistic regression also are consistent with our primary survival analysis findings that COVID-19 infection and Shingrix vaccination have independent effects on LOAD risk and that their non-additive joint effect also contributes significantly to LOAD risk. While holding Shingrix vaccination constant, the odds of LOAD are 16% higher among those with at least one reported COVID-19 infection compared to those without evidence of infection (OR=1.16, 95% CI: 1.13-1.18, p<0.0001) (**Figure 2c**). Conversely, while holding COVID-19 infection constant, the odds of LOAD are 16% lower among those with evidence of a Shingrix vaccination compared to those without (OR=0.84, 95% CI: 0.81-0.88, p<3.0x10^-15^) (**Figure 2c**). The COVID-19 and Shingrix results remained consistent with previous models, and the COVID-19 x Shingrix vaccination interaction term was statistically significant in this full model (OR=0.93; 95% CI: 0.88-0.98, p=0.009) (**Figure 2d**), indicating that the presence of COVID-19 infection modestly attenuates the protective effect of Shingrix vaccination on LOAD risk. This finding is consistent with the interaction term from the time-to-event analyses when both COVID-19 (HZ=1.14) and Shingrix vaccination (0.71) are present, resulting in an estimated joint HR of ∼0.955.

The association between Shingrix vaccination and LOAD may reflect true protection related to neuroinflammation or be a proxy for healthy behaviors, access to care, or general vaccine compliance. To test the latter possibilities, we captured information regarding a patient’s screening history, including mammography, colonoscopy, and prostate-specific antigen (PSA) procedures and tests, to serve as a proxy for individual health behaviors, as these preventive cancer screenings serve as indicators of health-seeking behavior, healthcare access, and adherence to recommended preventive care guidelines. We included if a patient has had at least one of these screenings as part of the propensity score matching criteria and re-ran the logistic regression models. Results remained unchanged when preventive healthcare services were included in matching. To examine whether vaccinations in general have a protective association with LOAD or if it is limited to vaccines that target herpes zoster, we tested for associations between LOAD and the pneumococcal vaccine, a vaccine recommended for the same age group and often administered alongside other routine vaccines. We found that, unlike Shingrix, the pneumococcal vaccine was not associated with LOAD when adjusted for age at baseline, sex, race, and number of clinic visits (OR=0.99, 95% CI: 0.97-1.01, p=0.35).

## Conclusions

Our results highlight the inverse impact of reported COVID-19 infection and Shingrix vaccination on LOAD risk and greatly extend recent findings on general dementia by examining how these two common exposures interact. A study focused on the recombinant zoster vaccine and based on EHRs available for analysis through the TriNetX U.S. Collaborative Network found that Shingrix associated with a 17% reduction in dementia risk over a six-year follow-up in over 200,000 patients^8^. An EHR study in Wales demonstrated the live attenuated herpes zoster vaccine reduced dementia incidence by approximately 20% over a seven-year follow-up^9^. Our study corroborates these protective effects specifically for LOAD, finding comparable if not somewhat larger effect sizes for Shingrix vaccination (27-33% reduction). Importantly, our analyses examined LOAD and its relationship not only with Shingrix vaccination but also with COVID-19, an infection associated with recurrence of shingles^28^ among other consequences^29^ of a suppressed immune state. The consistency of protective effects in three large-scale cohort studies, despite differences in study populations, study designs, and follow-up periods, strengthens the evidence for a protective association between Shingrix vaccination and neurodegenerative disease risk. Our results extend these findings by providing the first evidence that COVID-infection diminishes LOAD protection conferred by Shingrix vaccination. The associations described here cannot be explained by general patterns of healthy behaviors or healthcare utilization patterns, further implicating neuroinflammation as an important pathway in LOAD development.

This large-scale retrospective study of approximately 1.5 million patients found evidence that reported COVID-19 infection associated with increased incident LOAD risk, while Shingrix associated with lower incident LOAD risk. Multiple modeling strategies implemented here indicate that associations between COVID-19 and LOAD are consistent regardless of age at baseline, sex, race, and number of clinic visits, while the protective effect of Shingrix slightly decreases with age. Further, when both exposures were included in combined models, the effect estimates from time-fixed and time-varying models remained virtually unchanged, confirming that COVID-19 infection and Shingrix vaccination status represent independent risk and protective factors, respectively, for incident LOAD. The modestly significant interaction between the two exposures indicates that COVID-19 infection may decrease the protective effect of Shingrix, though the extent of the attenuation does not completely abolish the protective effects of the vaccination.

This study has several limitations and strengths. A limitation is our inability to include COVID vaccinations. COVID-19 vaccinations were not considered as vaccination status in the U.S. EHR suffers from substantial missingness. COVID-19 vaccinations relied heavily on mass vaccination sites, pharmacies, and drive-through clinics operating outside traditional healthcare settings, resulting in substantial lack of reporting within personal medical records compared to routine immunizations administered in clinical settings, such as Shingrix and pneumococcal vaccines. Also, while most older Americans did receive an initial dose^30,31^, many had already had a COVID-19 infection^17^. Finally, compared with a vaccination such as the sole recombinant zoster vaccination available in U.S. (Shingrix since 2017), several options were ultimately available for COVID-19, with evolving recommendations on booster updates and frequencies for both at-risk and general populations. While Shingrix vaccination status likely also suffers from similar missingness in this dataset (e.g., not always clinically captured or documented), the magnitude of protection against incident LOAD observed here is similar to that of a study in Wales with more complete medical records^9^.

An additional limitation is that the follow-up since baseline in this study is relatively short, necessarily constrained by the emergence of SARS-CoV-2/COVID-19 in the U.S. and the end of the N3C Data Transfer Agreements (initially designed for five years). Hence, the association between reported COVID-19 infection and the development of LOAD observed here may be an underestimate as more years of follow-up may reveal additional LOAD cases associated with COVID-19 infection, or alternatively, protective effects observed for interventions like Shingrix vaccination may attenuate over time. However, while follow-up in the present study is limited, it is the longest reported to date for comparable studies of COVID-19 infection and risk of dementia. One study using TriNetX examined a short window between February 2, 2020, and May 30, 2021, and estimated a higher Cox proportional hazards ratio for LOAD (1.69) within 360 days after COVID-19 infection^32^ compared with the wider window and lower hazards ratio (1.13) demonstrated here. Inflated hazards ratio may be due to reserve causality, overfitting, and immortal time bias among other causes. Differences in quality of phenotypes (e.g., single mention of single ICD-10 codes^32^ versus vetted concept sets^26^) and definition and inclusion of covariates may explain the differences in observed hazards ratios between this N3C study and the TriNetX study. Also, TriNetX does not deduplicate patient records as is required in N3C, and duplication of patient data could upwardly bias risk estimates when left uncorrected. The wider follow-up window considered here coupled with the multiple modeling strategies indicate the risk of incident LOAD is increased with COVID-19 infection but not at the magnitude previously suggested.

While this study took care in addressing confounding and several expected biases through propensity score matching and multiple modeling strategies, including time-vary exposure definitions, unmeasured confounding and other biases may still be present. Also, other characteristics of this pandemic, such as unrecognized infections and the rapidly changing SARS-CoV-2 variants, were not completely captured by the medical records underlying the N3C data resource. Although crude variant proxies such as year of infection are available, the high rate of reinfection makes it difficult to assess the impact of any one SARS-CoV-2 variant on the risk of LOAD.

A major strength of the study is sample size, which allowed for a variety of sensitivity analyses that examined potential biases such as survival bias and healthy behavior bias. With respect to the latter, we performed propensity score matching on age at baseline, sex, race, and preventive health behaviors (mammography, colonoscopy, or prostate-specific antigen screenings) that yielded similar results compared with the main analyses. The stability of the findings strongly indicate that our Shingrix associations represent vaccine-specific protection rather than representing a proxy for general health behaviors or vaccination compliance.

Protection from LOAD via Shingrix as opposed to healthy behaviors or vaccine uptake in general was further supported by the lack of association with the pneumococcal vaccine, a vaccine often administered alongside other routine vaccines. Recent reports suggest that protection from LOAD may be conferred by other AS01-adjuvantes like the respiratory syncytial virus (RSV) vaccine^33^, but further study is needed to distinguish between the impact on LOAD of the vaccines themselves or the infections they prevent.

Another major strength that the N3C infrastructure provides versus other data sources is the harmonization process of their housed EHR data. The N3C collected and harmonized EHR data from over 80 healthcare systems across the United States using the OMOP common data model to form final combined tables within the N3C data enclave^19,34^, thereby enhancing generalizability and reducing site-specific biases. Additionally, the concept set creation enabled the use of pre-established, validated definitions for various phenotypes and COVID-19 diagnoses specifically, to ensure consistency and reproducibility across different research efforts. While similar aggregated EHR resources like TriNetX and Epic Cosmos are larger than the N3C and likely overlap with N3C, both are proprietary systems and neither are freely available for research. Detailed documentation is also lacking for both TriNetX and Epic Cosmos whereas N3C provides clear, documented data governance and data standardization that follow the FAIR principles (Findable, Accessible, Interoperable, and Reusable^35^).

Overall, we identified two risk factors for incident LOAD among older individuals. First, avoidance of herpes zoster through vaccination is one strategy to decrease LOAD incidence in the long term. Second is the prevention of COVID-19 infection, although the effect of its vaccination is still to be determined.

## Supporting information

Supplementary Figure 1. Final National Clinical Cohort Collaborative (N3C) study population after exclusions.

Supplementary Figure 2. Covariate balance before and after propensity score matching.

## Data Availability

Code workbooks used are available upon request to the authors. Data are available to N3C level-2 certified users.

## Acknowledgements

This project was supported by the Clinical and Translational Science Collaborative of Northern Ohio which is funded by the National Center for Advancing Translational Sciences (NCATS) of the National Institutes of Health, UM1TR004528. The content is solely the responsibility of the authors and does not necessarily represent the official views of the NIH. This work was additionally supported by P30 AG072959, and CR was supported by T32 AG071474 and the AIM-AHEAD Consortium’s partnership with NCATS for a traineeship in Artificial Intelligence and Machine Learning. The analyses described in this publication were conducted with data or tools accessed through the NCATS N3C Data Enclave https://covid.cd2h.org and N3C Attribution & Publication Policy v 1.2-2020-08-25b supported by NCATS Contract No. 75N95023D00001, Axle Informatics Subcontract: NCATS-P00438-B. This research was possible because of the patients whose information is included within the data and the organizations (https://ncats.nih.gov/n3c/resources/data-contribution/data-transfer-agreement-signatories) and scientists who have contributed to the on-going development of this community resource [https://doi.org/10.1093/jamia/ocaa196]. The authors acknowledge the contribution of the N3C and the N3C Consortium members for making this research possible.

We gratefully acknowledge the following core contributors to N3C:

Adam B. Wilcox, Adam M. Lee, Alexis Graves, Alfred (Jerrod) Anzalone, Amin Manna, Amit Saha, Amy Olex, Andrea Zhou, Andrew E. Williams, Andrew M. Southerland, Andrew T. Girvin, Anita Walden, Anjali Sharathkumar, Benjamin Amor, Benjamin Bates, Brian Hendricks, Brijesh Patel, G. Caleb Alexander, Carolyn T. Bramante, Cavin Ward-Caviness, Charisse Madlock-Brown, Christine Suver, Christopher G. Chute, Christopher Dillon, Chunlei Wu, Clare Schmitt, Cliff Takemoto, Dan Housman, Davera Gabriel, David A. Eichmann, Diego Mazzotti, Donald E. Brown, Eilis Boudreau, Elaine L. Hill, Emily Carlson Marti, Emily R. Pfaff, Evan French, Farrukh M Koraishy, Federico Mariona, Fred Prior, George Sokos, Greg Martin, Harold P. Lehmann, Heidi Spratt, Hemalkumar B. Mehta, J.W. Awori Hayanga, Jami Pincavitch, Jaylyn Clark, Jeremy Richard Harper, Jessica Yasmine Islam, Jin Ge, Joel Gagnier, Johanna J. Loomba, John B. Buse, Jomol Mathew, Joni L. Rutter, Julie A. McMurry, Justin Guinney, Justin Starren, Karen Crowley, Katie Rebecca Bradwell, Kellie M. Walters, Ken Wilkins, Kenneth R. Gersing, Kenrick Cato, Kimberly Murray, Kristin Kostka, Lavance Northington, Lee Pyles, Lesley Cottrell, Lili M. Portilla, Mariam Deacy, Mark M. Bissell, Marshall Clark, Mary Emmett, Matvey B. Palchuk, Melissa A. Haendel, Meredith Adams, Meredith Temple-O’Connor, Michael G. Kurilla, Michele Morris, Nasia Safdar, Nicole Garbarini, Noha Sharafeldin, Ofer Sadan, Patricia A. Francis, Penny Wung Burgoon, Philip R.O. Payne, Randeep Jawa, Rebecca Erwin-Cohen, Rena C. Patel, Richard A. Moffitt, Richard L. Zhu, Rishikesan Kamaleswaran, Robert Hurley, Robert T. Miller, Saiju Pyarajan, Sam G. Michael, Samuel Bozzette, Sandeep K. Mallipattu, Satyanarayana Vedula, Scott Chapman, Shawn T. O’Neil, Soko Setoguchi, Stephanie S. Hong, Steven G. Johnson, Tellen D. Bennett, Tiffany J. Callahan, Umit Topaloglu, Valery Gordon, Vignesh Subbian, Warren A. Kibbe, Wenndy Hernandez, Will Beasley, Will Cooper, William Hillegass, Xiaohan Tanner Zhang. Details of contributions available at https://covid.cd2h.org/core-contributors

## Data Partners with Released Data

The following institutions whose data is released or pending:

Available: Advocate Health Care Network — UL1TR002389: The Institute for Translational Medicine (ITM) • Aurora Health Care Inc — UL1TR002373: Wisconsin Network For Health Research • Boston University Medical Campus — UL1TR001430: Boston University Clinical and Translational Science Institute • Brown University — U54GM115677: Advance Clinical Translational Research (Advance-CTR) • Carilion Clinic — UL1TR003015: iTHRIV Integrated Translational health Research Institute of Virginia • Case Western Reserve University — UL1TR002548: The Clinical & Translational Science Collaborative of Cleveland (CTSC) • Charleston Area Medical Center — U54GM104942: West Virginia Clinical and Translational Science Institute (WVCTSI) • Children’s Hospital Colorado — UL1TR002535: Colorado Clinical and Translational Sciences Institute • Columbia University Irving Medical Center — UL1TR001873: Irving Institute for Clinical and Translational Research • Dartmouth College — None (Voluntary) Duke University — UL1TR002553: Duke Clinical and Translational Science Institute • George Washington Children’s Research Institute — UL1TR001876: Clinical and Translational Science Institute at Children’s National (CTSA-CN) • George Washington University — UL1TR001876: Clinical and Translational Science Institute at Children’s National (CTSA-CN) • Harvard Medical School — UL1TR002541: Harvard Catalyst • Indiana University School of Medicine — UL1TR002529: Indiana Clinical and Translational Science Institute • Johns Hopkins University — UL1TR003098: Johns Hopkins Institute for Clinical and Translational Research • Louisiana Public Health Institute — None (Voluntary) • Loyola Medicine — Loyola University Medical Center • Loyola University Medical Center — UL1TR002389: The Institute for Translational Medicine (ITM) • Maine Medical Center — U54GM115516: Northern New England Clinical & Translational Research (NNE-CTR) Network • Mary Hitchcock Memorial Hospital & Dartmouth Hitchcock Clinic — None (Voluntary) • Massachusetts General Brigham — UL1TR002541: Harvard Catalyst • Mayo Clinic Rochester — UL1TR002377: Mayo Clinic Center for Clinical and Translational Science (CCaTS) • Medical University of South Carolina — UL1TR001450: South Carolina Clinical & Translational Research Institute (SCTR) • MITRE Corporation — None (Voluntary) • Montefiore Medical Center — UL1TR002556: Institute for Clinical and Translational Research at Einstein and Montefiore • Nemours — U54GM104941: Delaware CTR ACCEL Program • NorthShore University HealthSystem — UL1TR002389: The Institute for Translational Medicine (ITM) • Northwestern University at Chicago — UL1TR001422: Northwestern University Clinical and Translational Science Institute (NUCATS) • OCHIN — INV-018455: Bill and Melinda Gates Foundation grant to Sage Bionetworks • Oregon Health & Science University — UL1TR002369: Oregon Clinical and Translational Research Institute • Penn State Health Milton S. Hershey Medical Center — UL1TR002014: Penn State Clinical and Translational Science Institute • Rush University Medical Center — UL1TR002389: The Institute for Translational Medicine (ITM) • Rutgers, The State University of New Jersey — UL1TR003017: New Jersey Alliance for Clinical and Translational Science • Stony Brook University — U24TR002306 • The Alliance at the University of Puerto Rico, Medical Sciences Campus — U54GM133807: Hispanic Alliance for Clinical and Translational Research (The Alliance) • The Ohio State University — UL1TR002733: Center for Clinical and Translational Science • The State University of New York at Buffalo — UL1TR001412: Clinical and Translational Science Institute • The University of Chicago — UL1TR002389: The Institute for Translational Medicine (ITM) • The University of Iowa — UL1TR002537: Institute for Clinical and Translational Science • The University of Miami Leonard M. Miller School of Medicine — UL1TR002736: University of Miami Clinical and Translational Science Institute • The University of Michigan at Ann Arbor — UL1TR002240: Michigan Institute for Clinical and Health Research • The University of Texas Health Science Center at Houston — UL1TR003167: Center for Clinical and Translational Sciences (CCTS) • The University of Texas Medical Branch at Galveston — UL1TR001439: The Institute for Translational Sciences • The University of Utah — UL1TR002538: Uhealth Center for Clinical and Translational Science • Tufts Medical Center — UL1TR002544: Tufts Clinical and Translational Science Institute • Tulane University — UL1TR003096: Center for Clinical and Translational Science • The Queens Medical Center — None (Voluntary) • University Medical Center New Orleans — U54GM104940: Louisiana Clinical and Translational Science (LA CaTS) Center • University of Alabama at Birmingham — UL1TR003096: Center for Clinical and Translational Science • University of Arkansas for Medical Sciences — UL1TR003107: UAMS Translational Research Institute • University of Cincinnati — UL1TR001425: Center for Clinical and Translational Science and Training • University of Colorado Denver, Anschutz Medical Campus — UL1TR002535: Colorado Clinical and Translational Sciences Institute • University of Illinois at Chicago — UL1TR002003: UIC Center for Clinical and Translational Science • University of Kansas Medical Center — UL1TR002366: Frontiers: University of Kansas Clinical and Translational Science Institute • University of Kentucky — UL1TR001998: UK Center for Clinical and Translational Science • University of Massachusetts Medical School Worcester — UL1TR001453: The UMass Center for Clinical and Translational Science (UMCCTS) • University Medical Center of Southern Nevada — None (voluntary) • University of Minnesota — UL1TR002494: Clinical and Translational Science Institute • University of Mississippi Medical Center — U54GM115428: Mississippi Center for Clinical and Translational Research (CCTR) • University of Nebraska Medical Center — U54GM115458: Great Plains IDeA-Clinical & Translational Research • University of North Carolina at Chapel Hill — UL1TR002489: North Carolina Translational and Clinical Science Institute • University of Oklahoma Health Sciences Center — U54GM104938: Oklahoma Clinical and Translational Science Institute (OCTSI) • University of Pittsburgh — UL1TR001857: The Clinical and Translational Science Institute (CTSI) • University of Pennsylvania — UL1TR001878: Institute for Translational Medicine and Therapeutics • University of Rochester — UL1TR002001: UR Clinical & Translational Science Institute • University of Southern California — UL1TR001855: The Southern California Clinical and Translational Science Institute (SC CTSI) • University of Vermont — U54GM115516: Northern New England Clinical & Translational Research (NNE-CTR) Network • University of Virginia — UL1TR003015: iTHRIV Integrated Translational health Research Institute of Virginia • University of Washington — UL1TR002319: Institute of Translational Health Sciences • University of Wisconsin-Madison — UL1TR002373: UW Institute for Clinical and Translational Research • Vanderbilt University Medical Center — UL1TR002243: Vanderbilt Institute for Clinical and Translational Research • Virginia Commonwealth University — UL1TR002649: C. Kenneth and Dianne Wright Center for Clinical and Translational Research • Wake Forest University Health Sciences — UL1TR001420: Wake Forest Clinical and Translational Science Institute • Washington University in St. Louis — UL1TR002345: Institute of Clinical and Translational Sciences • Weill Medical College of Cornell University — UL1TR002384: Weill Cornell Medicine Clinical and Translational Science Center • West Virginia University — U54GM104942: West Virginia Clinical and Translational Science Institute (WVCTSI) Submitted: Icahn School of Medicine at Mount Sinai — UL1TR001433: ConduITS Institute for Translational Sciences • The University of Texas Health Science Center at Tyler — UL1TR003167: Center for Clinical and Translational Sciences (CCTS) • University of California, Davis — UL1TR001860: UCDavis Health Clinical and Translational Science Center • University of California, Irvine — UL1TR001414: The UC Irvine Institute for Clinical and Translational Science (ICTS) • University of California, Los Angeles — UL1TR001881: UCLA Clinical Translational Science Institute • University of California, San Diego — UL1TR001442: Altman Clinical and Translational Research Institute • University of California, San Francisco — UL1TR001872: UCSF Clinical and Translational Science Institute  NYU Langone Health Clinical Science Core, Data Resource Core, and PASC Biorepository Core — OTA-21-015A: Post-Acute Sequelae of SARS-CoV-2 Infection Initiative (RECOVER)  Pending: Arkansas Children’s Hospital — UL1TR003107: UAMS Translational Research Institute • Baylor College of Medicine — None (Voluntary) • Children’s Hospital of Philadelphia — UL1TR001878: Institute for Translational Medicine and Therapeutics • Cincinnati Children’s Hospital Medical Center — UL1TR001425: Center for Clinical and Translational Science and Training • Emory University — UL1TR002378: Georgia Clinical and Translational Science Alliance • HonorHealth — None (Voluntary) • Loyola University Chicago — UL1TR002389: The Institute for Translational Medicine (ITM) • Medical College of Wisconsin — UL1TR001436: Clinical and Translational Science Institute of Southeast Wisconsin • MedStar Health Research Institute — None (Voluntary) • Georgetown University — UL1TR001409: The Georgetown-Howard Universities Center for Clinical and Translational Science (GHUCCTS) • MetroHealth — None (Voluntary) • Montana State University — U54GM115371: American Indian/Alaska Native CTR • NYU Langone Medical Center — UL1TR001445: Langone Health’s Clinical and Translational Science Institute • Ochsner Medical Center — U54GM104940: Louisiana Clinical and Translational Science (LA CaTS) Center • Regenstrief Institute — UL1TR002529: Indiana Clinical and Translational Science Institute • Sanford Research — None (Voluntary) • Stanford University — UL1TR003142: Spectrum: The Stanford Center for Clinical and Translational Research and Education • The Rockefeller University — UL1TR001866: Center for Clinical and Translational Science • The Scripps Research Institute — UL1TR002550: Scripps Research Translational Institute • University of Florida — UL1TR001427: UF Clinical and Translational Science Institute • University of New Mexico Health Sciences Center — UL1TR001449: University of New Mexico Clinical and Translational Science Center • University of Texas Health Science Center at San Antonio — UL1TR002645: Institute for Integration of Medicine and Science • Yale New Haven Hospital — UL1TR001863: Yale Center for Clinical Investigation

## Disclaimer

The N3C Publication committee confirmed that this manuscript MSID: 2728.417 is in accordance with N3C data use and attribution policies; however, this content is solely the responsibility of the authors and does not necessarily represent the official views of the National Institutes of Health or the N3C program.

