## Supplementary Figure 1. Final National Clinical Cohort Collaborative (N3C) study population after exclusions. for "Opposing and antagonizing effects of SARS-CoV-2/COVID-19 infection and recombinant zoster vaccination on the risk of late-onset Alzheimer disease"

**Supplementary Figure 1. Final National Clinical Cohort Collaborative (N3C) study population after exclusions.** The study period was January 20, 2020 (date of first reported US confirmed COVID-19 case), to May 16, 2025 (date of final N3C data release for COVID-19 tenant). Patients were excluded if they had a zero-visit history in the defined baseline period (January 20, 2019, and January 20, 2020), had an LOAD diagnosis before the baseline date (January 20, 2020), had a mention of early onset AD, had missingness for race or sex, entry errors for COVID-19 or Shingrix vaccination, and/or were under the age of 65.

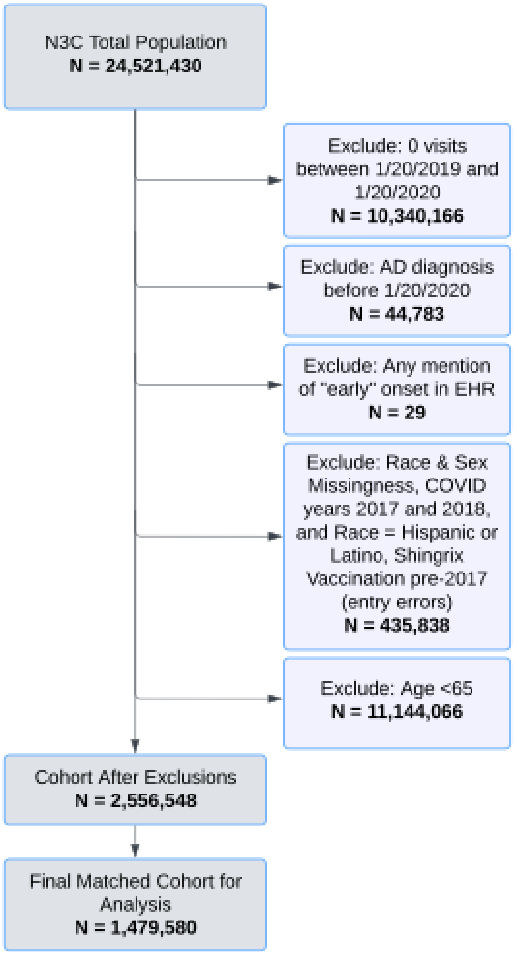
