## Supplementary Figure 2. Covariate balance before and after propensity score matching. for "Opposing and antagonizing effects of SARS-CoV-2/COVID-19 infection and recombinant zoster vaccination on the risk of late-onset Alzheimer disease"

**Supplementary Figure 2. Covariate balance before and after propensity score matching.** Propensity score matching was performed using the *matchit* R package and 1:1 nearest neighbor methods to ensure COVID-19-confirmed cases and non-cases have similar distributions based on age, sex, and race. Plotted are the absolute standardized mean differences (x-axis) for each for the covariates (y-axis) before matching (open circles) and after matching (closed circles) on the exposure of interest (COVID-19 infection). The dashed line at 0.10 represents the conventional balance threshold. Shown are Love plots for the a) main matching equation and b) matching equation with the health services variable included.

**a)**

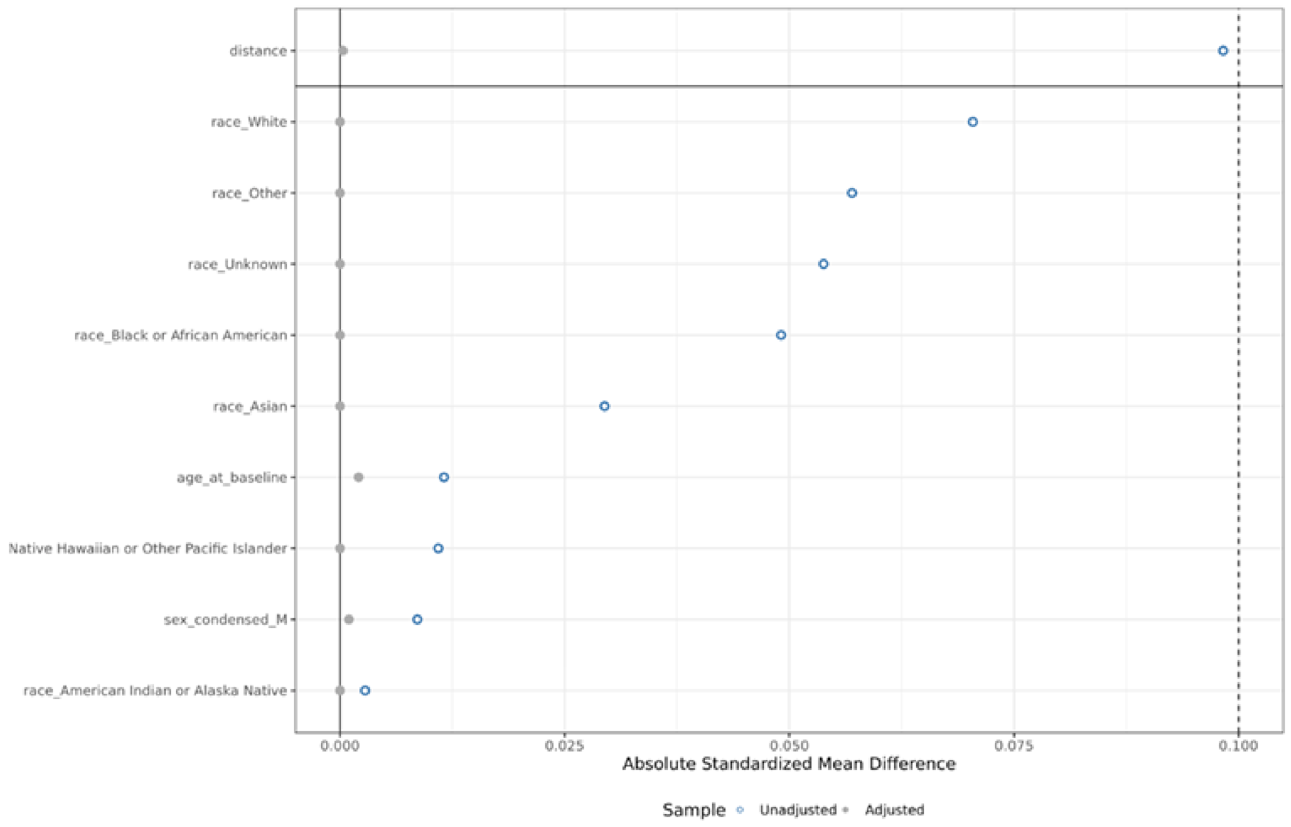

**b)**

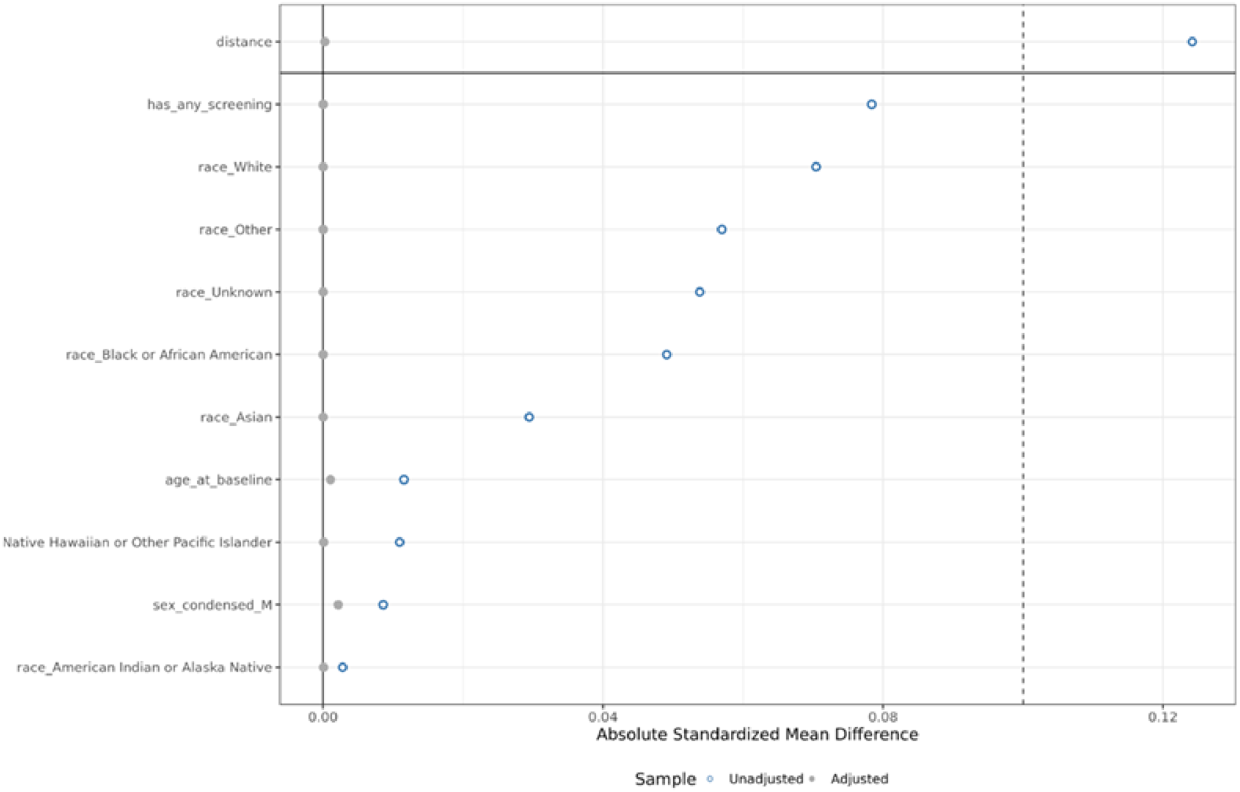
distance
has_any_screening
race_White
race_Other
race_Unknown
race_Black or African American
race_Asian
age_at_baseline
Native Hawaiian or Other Pacific Islander
sex_condensed_M
race_American Indian or Alaska Native
0.00
0.04
0.08
0.12
Absolute Standardized Mean Difference
Sample
Unadjusted
Adjusted
